# Sex differences in innate anti-viral immune responses to respiratory viruses

**DOI:** 10.1101/2020.09.18.20195784

**Authors:** Eteri Regis, Sara Fontanella, Lijing Lin, Rebecca Howard, Sadia Haider, John A. Curtin, Michael R. Edwards, Magnus Rattray, Angela Simpson, Adnan Custovic, Sebastian L. Johnston

## Abstract

Males have excess morbidity and mortality from respiratory viral infections and especially so in COVID-19. The mechanisms explaining this excess in disease burden in males are unknown. Innate immune responses are likely critical in protection against a novel virus like SARS-CoV-2. We hypothesised that innate immune responses may be deficient in males relative to females. To test this we stimulated peripheral blood mononuclear cells (PBMCs) from participants in a population-based birth cohort with three respiratory viruses (rhinoviruses-RV-A16 and RV-A1, and respiratory syncytial virus-RSV) and two viral mimics (R848 and CpG-A, to mimic responses to SARS-CoV-2). We measured interferon (IFN) and IFN-induced chemokine responses and investigated sex differences in virus-induced responses. IFN-α, IFN-β and IFN-γ responses to RV-A16 were deficient in males compared to females, fold-inductions being 1.92-fold (*P<*0.001), 2.04-fold (*P*<0.001) and 1.77-fold (*P*=0.003) lower in males than females, respectively. Similar significant deficiencies in innate immune responses were observed in males for eleven other cytokine-stimulus pairs. Responses in males were greater than those in females in only one of 35 cytokine-stimulus pairs investigated. Review of healthcare records revealed that 12.1% of males but only 6.6% of females were admitted to hospital with respiratory infections in the first year of life (*P*=0.017). Impaired innate anti-viral immunity in males likely results in high morbidity and mortality from respiratory viruses including COVID-19. Males may preferentially benefit from therapies that boost innate anti-viral immune responses.

**Significance Statement:** Clinical outcomes including, mortality, Intensive care unit admissions and hospital admissions, during COVID-19 disease are consistently and substantially worse in males than in females. The mechanisms underlying this increased susceptibility to severe disease in males are not understood. We hypothesised that the differential outcomes between sexes could be a consequence of deficient innate interferon responses in males, and more robust innate interferon responses in females. We have investigated such responses in a large population-based cohort and found that indeed males have deficient innate interferon responses to viral stimuli, including stimuli that mimic SARS-CoV-2 infection, relative to females. Our findings have very important treatment implications as interferons are available for clinical use with immediate effect.

## Introduction

Respiratory viral infections are a leading cause of ill health and mortality(1). This has been highlighted by the immense global impact of COVID-19(2) and the enormous public health threat it poses. Males and females differ in the prevalence and severity of viral infections(3). In COVID-19, risk factors for mortality include older age, the presence of comorbid conditions, and male sex(4, 5). In an early report of 191 cases from Wuhan, China, 70% of deaths were male, while only 30% were female(5). This gender imbalance has subsequently been confirmed in a large study of 44,672 COVID-19 confirmed cases in China, where 63.8% of deaths were male while only 36.2% were female, and the case fatality rate for males was 2.8%, while that for females was 1.7%(6). Similar mortality data are reported in Italy where 9390 of 13,334 (70.4%) of COVID-19 deaths in persons under 90 years of age reported by 6^th^ April 2020 were male, and 3,944 (29.6%) were female(7). In the UK, the data are very similar, with males representing 65% of deaths involving COVID-19 reported by 27^th^ March 2020, while 35% were females(8). Intensive Care Unit (ICU) admission are also much higher for males in Lombardy, Italy, where 82% of 1591 ICU admissions were male(9). The ICU data are very similar in the UK, as 72.5% of 3883 confirmed COVID-19 cases admitted to ICU by the 10^th^ April 2020 were male, and 27.5% female(10). A recent large UK study of 20,133 patients admitted to hospital with COVID-19, reported more men were admitted than women (men 60%, n=12 068; women 40%, n=8065)(11). An even larger study of primary care records of 17,278,392 adults linked to 10,926 COVID-19-related deaths reported COVID-19-related death was associated with: being male with a hazard ratio of 1.59(12). COVID-19 case identification in population screening also reports a male preponderance, with more males than females testing positive in both targeted testing (16.7% vs 11.0%) and in population screening (0.9% vs 0.6%)(13).

The mechanisms explaining excess COVID-19 mortality, ICU and hospital admissions and case identification in males are unknown(14). Understanding how sexes differ in their responses to respiratory viruses is critically important for the development of treatment and preventative strategies, which may differ for males and females. Innate immune responses, mediated by anti-viral interferon (IFN) production by virus-infected cells will be critical in protection against SARS-CoV-2, a new virus that humans have never previously encountered. We have previously reported that deficient IFN-α(15), IFN-β(15, 16) and IFN-γ(17, 18) production by virus-infected cells from people with asthma is implicated in their increased susceptibility to respiratory virus infections(19). However, little is known about the variation within the human immune system in relation to the patterns of response to viruses at a population level. We hypothesised that the adverse outcomes for males reported in COVID-19 may be related to deficient innate immune responses to viruses in males relative to females. To investigate this, we stimulated peripheral blood mononuclear cells (PBMCs) from male and female participants in a population-based birth cohort with three common respiratory viruses and two viral mimics (R848 and CpG-A, to mimic responses to SARS-CoV-2) and measured IFN and IFN-induced chemokine responses in supernatants.

## Results

### Participant flow and demographic data

Of 751 participants who attended follow-up at age 16 years, 361 provided blood samples. After quality control (Supplementary Appendix), we excluded data for 16 participants. Participant flow is presented in Fig. 1. There were no differences in demographic characteristics, environmental exposures and clinical features between participants included in this analysis (n=345) and those who were not (n=406), either in the whole population or stratified by sex (Table S1).

**Fig. 1:**
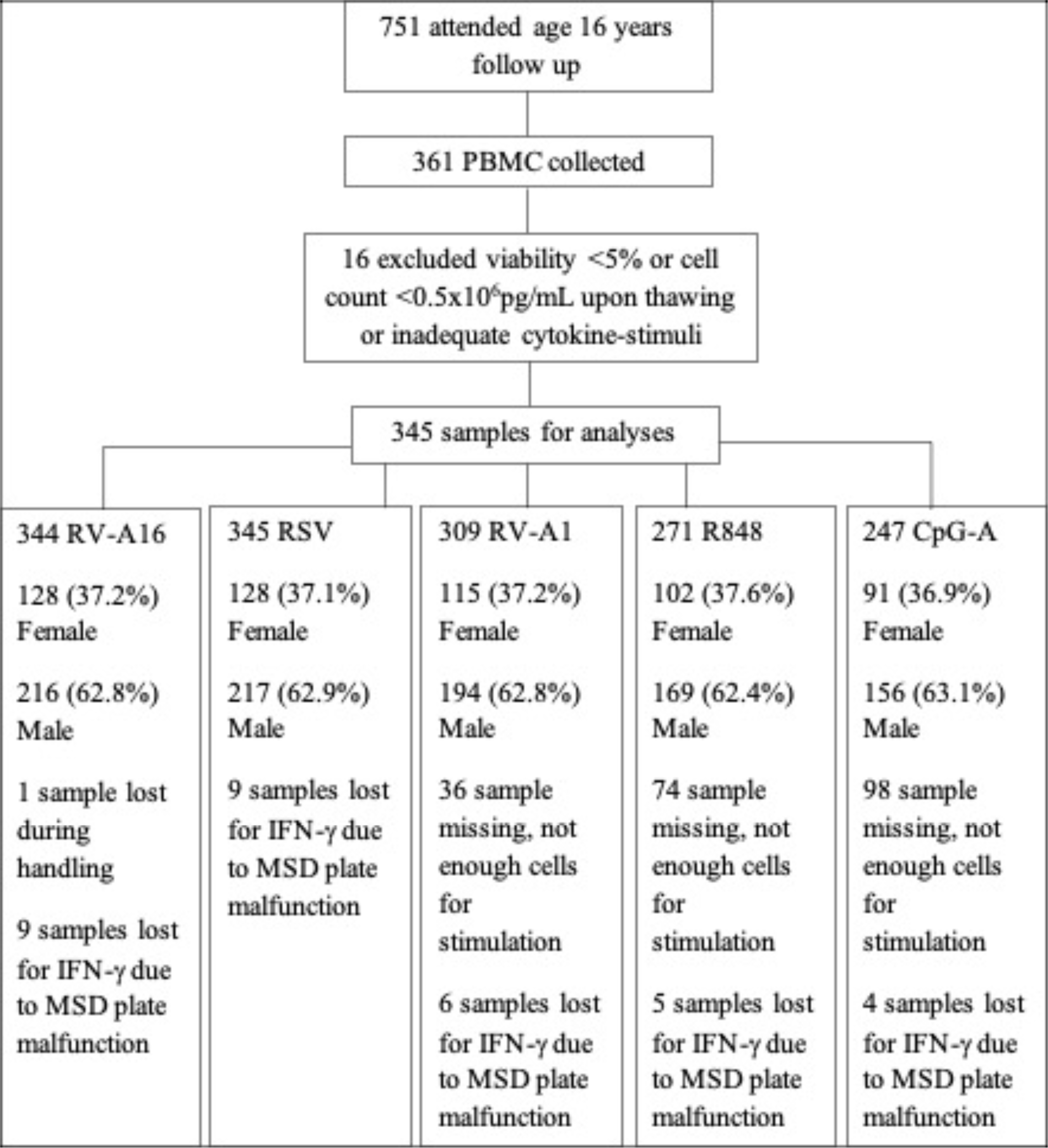
Participant flow and PBMC stimulation numbers for each stimulus.

The demographic and clinical characteristics of the 345 subjects included in this analysis are shown in Table 1. There were no significant differences between sexes in birth weight, relevant environmental exposures (including position in sibship, pet ownership and tobacco smoke exposure) or common respiratory diseases, such as wheezing and asthma. No significant differences were observed in cell viability between sexes (Fig. S1).

**Table 1:** Comparison between sexes of demographic and clinical characteristics of the study population. Some data was lost due to individuals not attending a follow up or skipping the question

|  | Cytokine data (n=345) |  |  |
| --- | --- | --- | --- |
| | Female<br>n (%) | Male<br>n (%) | p-value<br>$\chi^2$ |
| Ethnicity (Caucasian) | 121/126(96.0) | 204/212(96.2) | 0.504 |
| Younger siblings | 112/217(51.6) | 70/128(54.7) | 0.581 |
| Older siblings | 62/128 (48.4) | 121/215 (56.3) | 0.159 |
| Day care attendance | 91/121(75.2) | 153/209(73.2) | 0.690 |
| Maternal smoking (pregnancy) | 9/128 (7.0) | 19/212 (9.0) | 0.530 |
| Maternal smoking (current) | 10/126(7.9) | 29/217(13.4) | 0.127 |
| Maternal asthma | 23/128 (18.0) | 39/217 (18.0) | 0.999 |
| Paternal asthma | 22/128 (17.2) | 29/217 (13.4) | 0.334 |
| Dog ownership | 40/127(31.5) | 80/216(37.0) | 0.298 |
| Cat ownership | 29/127(22.8) | 54/215(25.1) | 0.634 |
| Current asthma | 20/126 (18.3) | 39/213 (18.3) | 0.567 |
| Current wheeze | 18/126(14.3) | 33/215 (15.3) | 0.790 |
|  | **Mean(SD)** | **Mean(SD)** | **t-test** |
| Age at follow up | 16.0 (0.65) | 16.1 (0.55) | 0.763 |
| Birth weight (kg) | 3.42 (0.47) | 3.51(0.91) | 0.242 |

### Induction of IFNs and IFN-induced chemokines in PBMCs in response to viral stimuli

There was a significant induction of all IFNs and IFN-induced chemokines in response to all viral stimuli compared to medium control (Figures 2 and S2). Cytokine responses to viral stimuli were not normally distributed (Shapiro-Wilk test, Table S2). The most potent inducers of IFN-α were CpG-A, RSV and RV-A16, with median concentrations of 184.7pg/mL, 108.3pg/mL and 34.0pg/mL respectively compared to 0.0pg/mL in medium control (all *P*<0.001, Figure 2). RV-A1 and R848 also induced IFN-α, but to lesser degrees (Fig. 2). Induction of IFN-β followed a very similar pattern to that of IFN-α, but with lower concentrations.

**Figure 2.**
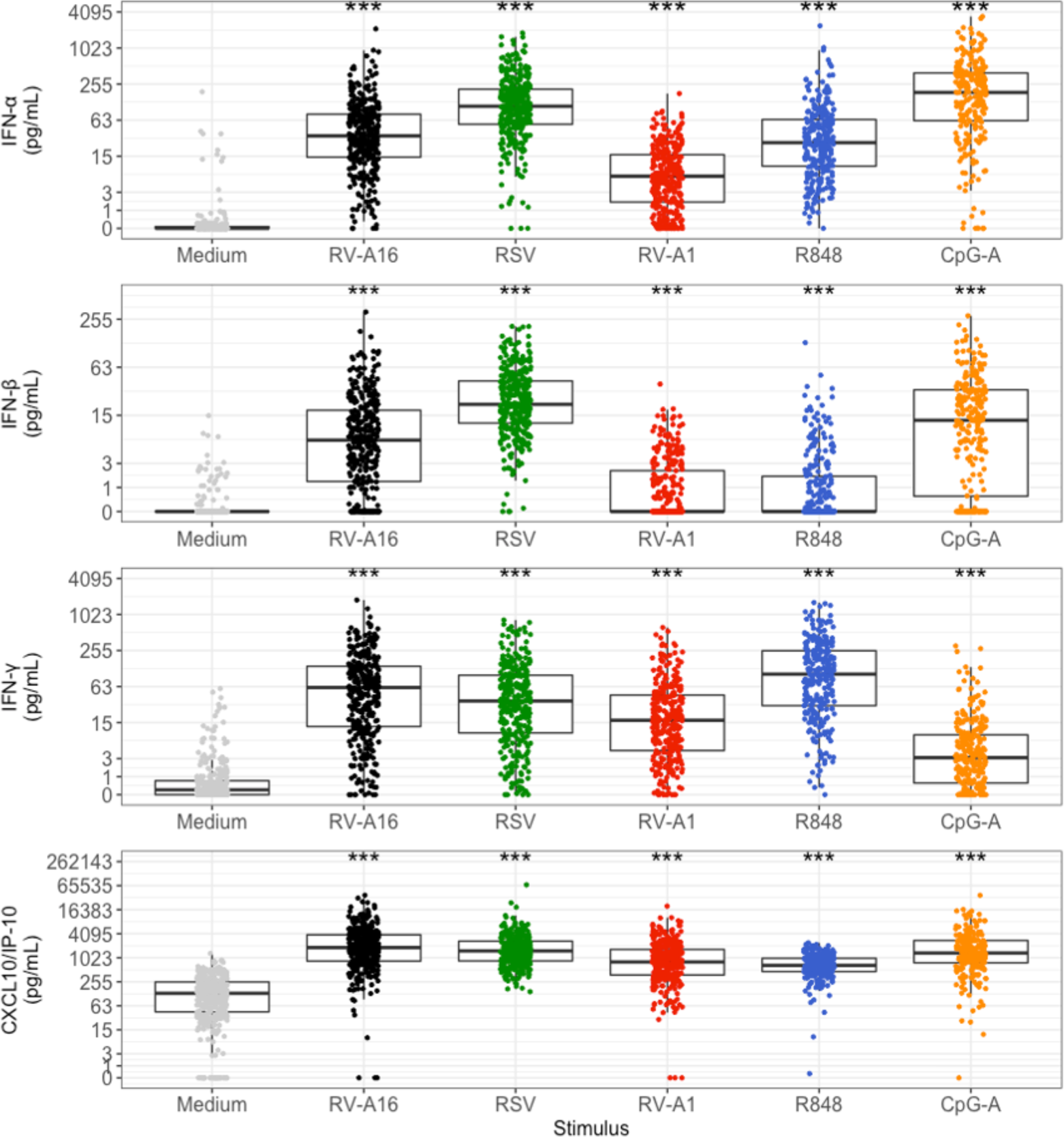
Patterns of PBMC cytokine induction by viral stimuli, compared to medium controls. Data were analysed using Wilcoxon test. Each dot represents an individual participant. Box plots represent the 25^th^ and 75^th^ percentiles, the line the median, with whiskers at the 10^th^ and 90^th^ percentiles. Data are presented in pg/mL. The y axis is plotted on a logarithmic scale. Significant levels: ^***^*P*<001 compared to medium.

The most potent inducers of IFN-γ were R848, RV-A16 and RSV, with median concentrations of 102.1pg/mL, 60.7pg/mL and 35.7pg/mL, respectively, compared to 0.2pg/mL in medium control (all *P*<0.001, Figure 2). RV-A1 and CpG-A also induced IFN-γ, but to lesser degrees (Fig. 2).

The most potent inducers of the IFN-induced chemokine CXCL10/IP-10, were RV-A16, RSV and CpG-A, with median concentrations of 1841.0pg/mL, 1515.2pg/mL and 1343.4pg/mL respectively, compared to 130.6pg/mL in medium control samples (all *P*<0.001, Figure 2). RV-A1 and R848 also induced CXCL-10/IP-10, but to lesser degrees.

The IFN-induced chemokines CCL2/MCP1, CCL4/MIP-1β and CCL13/MCP4 were also all induced by all viral stimuli, CCL2/MCP1 and CCL4/MIP-1β most potently by R848 and CCL13/MCP4 most potently by RV-A1 (Fig. S2).

### Differences between males and females in IFN and IFN-induced chemokine induction

Results of the comparisons between sexes are presented in Table 2. The trend across all viral stimuli and responses was remarkably consistent, as almost all of the IFNs and IFN-induced chemokines had higher levels of induction in females compared to males: out of 35 stimulus-cytokine pairs, females exhibited higher induction than males in 32 cases (Table 2).

**Table 2:** Differences between males and females in IFN and IFN-induced chemokine responses to viral stimuli. Data were analysed using the Wilcoxon test. *P* values and adjusted *P* values less than 0.05 are in bold. The group with the higher IFN and IFN-induced chemokine induction is highlighted in grey.

| Stimulus | Cytokine | Median log2 fold Induction [IQR] | Median log2 fold Induction [IQR] | Female to Male ratio in fold induction | *P* value | Adjusted *P* value |
| --- | --- | --- | --- | --- | --- | --- |
| RV-A16 |  | **Female, n=128 (37.2%)** | **Male, n=216 (62.8%)** |  |  |  |
| | IFN-α | 10.24 [8.98-11.55] | 9.30 [7.88-10.53] | 1.92 | **<0.001** | **0.001** |
| | IFN-β | 6.01 [4.66-7.26] | 4.98 [2.02-6.66] | 2.04 | **<0.001** | **0.003** |
| | IFN-γ | 7.69 [6.04-8.90] | 6.87 [5.03-8.15] | 1.77 | **0.003** | **0.017** |
| | CCL4/MIP-1β | 2.13 [1.47-3.25] | 1.98 [0.96-2.98] | 1.11 | 0.125 | 0.231 |
|  | CXCL10/IP-10 | 4.15 [3.13-5.27] | 3.66 [2.36-4.95] | 1.04 | **0.010** | **0.041** |
|  | CCL13/MCP4 | 2.60 [1.19-3.85] | 2.10 [0.93-3.41] | 1.41 | 0.076 | 0.149 |
|  | CCL2/MCP1 | 2.09 [1.09-2.95] | 1.91 [0.95-2.92] | 1.13 | 0.659 | 0.721 |
| RSV |  | **Female n=128 (37.1%)** | **Male n=217 (62.9%)** |  |  |  |
| | IFN-α | 11.66 [10.53-12.53] | 11.24 [9.91-12.21] | 1.34 | **0.018** | 0.055 |
| | IFN-β | 6.89 [5.83-7.49] | 6.62 [5.58-7.71] | 1.21 | 0.583 | 0.658 |
| | IFN-γ | 7.08 [5.88-8.14] | 6.54 [4.84-8.11] | 1.45 | 0.062 | 0.142 |
| | CCL4/MIP-1β | 1.60 [1.08-2.36] | 1.58 [0.98-2.20] | 1.01 | 0.380 | 0.475 |
|  | CXCL10/IP-10 | 3.67 [2.59-4.83] | 3.36 [2.38-4.62] | 1.24 | 0.210 | 0.304 |
|  | CCL13/MCP4 | 1.46 [0.42-2.51] | 1.12 [-0.11-2.13] | 1.27 | **0.019** | 0.055 |
|  | CCL2/MCP1 | 1.75 [0.93-2.53] | 1.64 [0.64-2.65] | 1.08 | 0.799 | 0.848 |
| RV-A1 |  | **Female n=115 (37.2%)** | **Male n=194 (62.8%)** |  |  |  |
| | IFN-α | 7.54 [5.68-8.92] | 6.91 [4.92-8.34] | 1.55 | **0.011** | **0.041** |
| | IFN-β | 1.21 [1.21-3.61] | 1.21 [1.21-3.16] | 1.00 | 0.483 | 0.583 |
| | IFN-γ | 6.08 [4.32-7.27] | 5.51 [3.69-6.83] | 1.48 | 0.066 | 0.142 |
| | CCL4/MIP-1β | 0.93 [0.43-1.41] | 0.71 [0.20-1.13] | 1.16 | **0.010** | **0.041** |
|  | CXCL10/IP-10 | 3.05 [1.94-4.17] | 2.75 [1.36-3.85] | 1.23 | **0.041** | 0.104 |
|  | CCL13/MCP4 | 3.39 [2.07-4.47] | 2.70 [0.67-3.80] | 1.61 | **0.002** | **0.017** |
|  | CCL2/MCP1 | 2.00 [1.09-2.77] | 1.84 [0.85-2.58] | 1.12 | 0.217 | 0.304 |
| R848 |  | **Female n=102 (37.6%)** | **Male n=169 (62.4%)** |  |  |  |
| | IFN-α | 9.91 [8.41-11.52] | 8.87 [7.47-10.26] | 2.06 | **<0.001** | **0.002** |
| | IFN-β | 1.53 [1.53-3.27] | 1.53 [1.53-2.84] | 1.00 | 0.158 | 0.261 |
| | IFN-γ | 7.51 [5.84-9.14] | 7.11 [5.45-8.60] | 1.32 | 0.164 | 0.261 |
| | CCL4/MIP-1β | 3.58 [2.94-4.45] | 3.46 [2.56-4.42] | 1.09 | 0.375 | 0.475 |
|  | CXCL10/IP-10 | 2.15 [1.25-3.15] | 1.79 [1.04-2.82] | 1.28 | 0.069 | 0.142 |
|  | CCL13/MCP4 | 0.83 [-0.31-1.85] | 0.76 [-0.85-1.69] | 1.05 | 0.239 | 0.322 |
|  | CCL2/MCP1 | 2.26 [0.79-3.14] | 2.02 [0.97-3.04] | 1.18 | 0.914 | 0.914 |
| CpG-A |  | **Female n=91 (36.9%)** | **Male n=156 (63.1%)** |  |  |  |
| | IFN-α | 12.6 [10.99-13.73] | 11.83 [10.47-13.38] | 1.71 | **0.015** | 0.051 |
| | IFN-β | 6.31 [2.86-8.10] | 5.71 [1.40-7.56] | 1.52 | 0.155 | 0.261 |
| | IFN-γ | 4.04 [2.29-5.17] | 3.03 [1.54-4.26] | 2.01 | **0.003** | **0.017** |
| | CCL4/MIP-1β | 1.35 [0.91-1.91] | 1.43 [0.87-1.81] | 0.95 | 0.857 | 0.882 |
|  | CXCL10/IP-10 | 4.02 [2.94-4.93] | 3.66 [2.71-4.67] | 1.28 | 0.204 | 0.304 |
|  | CCL13/MCP4 | 2.56 [1.70-3.53] | 2.12 [0.88-3.32] | 1.36 | **0.040** | 0.104 |
|  | CCL2/MCP1 | 1.66 [1.07-2.19] | 1.54 [0.89-2.07] | 1.09 | 0.528 | 0.616 |

The induction of IFN-α was significantly higher in females than in males for all five viral stimuli (with *P*-values ranging from <0.001 to 0.018, Table 2). After adjustment for multiple testing, these differences remained statistically significant for RV-A16, RV-A1 and R848 (*P*=0.001, 0.041 and 0.002 respectively), and marginal for RSV and CpG-A (*P*=0.055 and 0.051 respectively, Table 2). The magnitude of the differences observed for IFN-α ranged from a 1.34-fold (34%) greater induction of IFN-α in females than in males for RSV, to a 2.06-fold (106%) greater induction for R848.

The individual responses of each IFN and of CXCL10/IP-10 to RV-A16 stimulation are depicted in Fig. 3 A-D, with induction in females significantly greater than that in males for IFN-α at 1.92-fold (92%) greater (*P*<0.001), IFN-β at 2.04-fold (104%) greater (*P<*0.001), IFN-γ 1.77-fold (77%) (*P*=0.003) and CXCL10/IP-10 1.40-fold (40%) greater *(P*=0.01).

**Figure 3:**
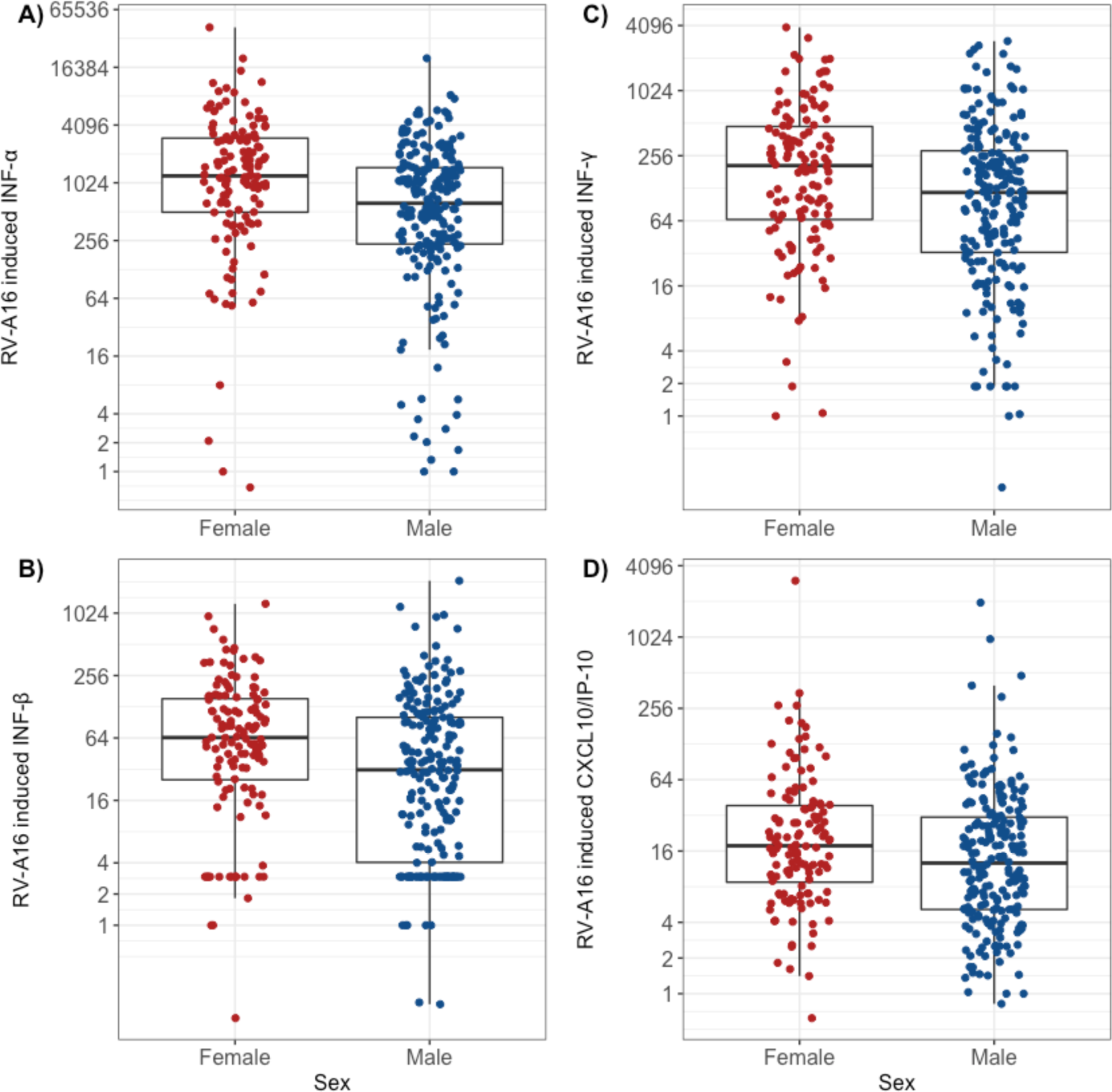
Females have significantly greater induction than males of type I and II IFNs and CXCL10/IP-10 in response to RV-A16. Box plots represent the 25^th^ and 75^th^ percentiles, the line the median, with whiskers at the 10^th^ and 90^th^ percentiles. Each dot represents an individual participant. Wilcoxon test: A) *P*<0.001, B) *P*<0.001, C) *P*=0.003 and D) *P*=0.010. Data are presented as fold induction. The y axis is plotted on a logarithmic scale.

Regarding the viral mimics R848 and CpG-A (which mimic responses to SARS-CoV-2 infection), females had significantly greater induction of IFN-α in response to R848 stimulation (2.06-fold, *P*<0.001), and in response to CpG-A (1.71-fold, *P*=0.015) (Fig. 4 A-B), though the latter difference was marginal after adjustment (*P*=0.051, Table 2). Females also had significantly greater induction of IFN-γ (2.01-fold, *P*=0.003) and CCL13/MCP4 (1.36-fold, *P*=0.04) in response to CpG-A than males (Fig. 4 C-D), though the latter difference was marginal after adjustment (*P*=0.10, Table 2).

**Figure 4:**
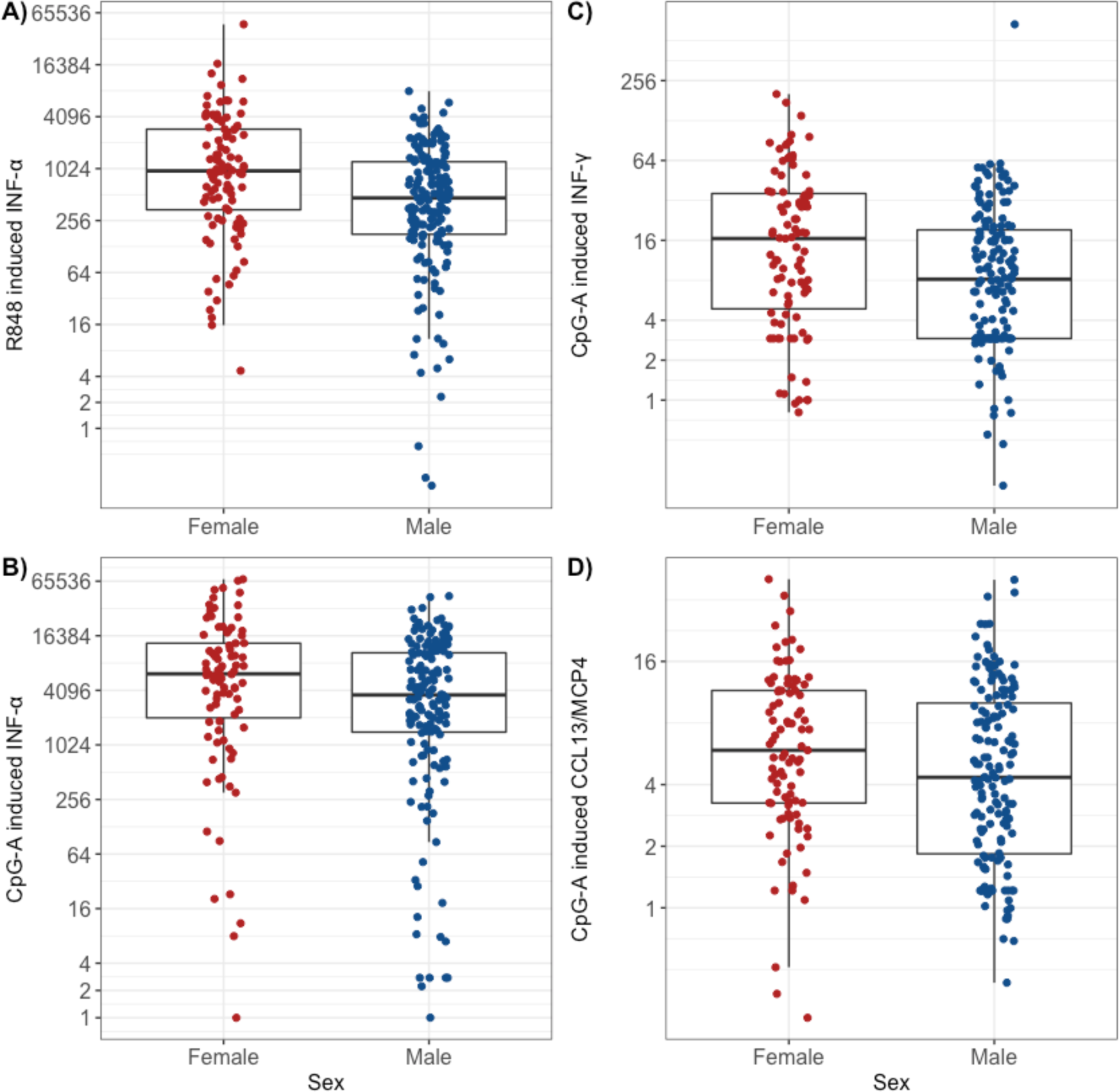
Females have significantly greater induction than males of type I and II IFNs and CCL13/MCP4 in response to the viral mimics R848 and CpG-A. Box plots represent the 25^th^ and 75^th^ percentiles, the line the median, with whiskers at the 10^th^ and 90^th^ precentiles. Each dot represents an individual participant. Wilcoxon test: A) *P*<0.001, B) *P*=0.015, C) *P*=0.003 and D) *P*=0.040. Data are presented as fold induction. The y axis is plotted on a logarithmic scale.

Stimulation with RV-A1 also resulted in significantly greater induction in females than in males for IFN-α (Fig. S3A, 1.55-fold, *P*=0.011), CXCL10/IP-10 (Figure S3B, 1.23-fold, *P*=0.041, though this difference was marginal after adjustment [*P*=0.10, Table 2]), CCL4/MIP-1β (Fig. S3C, 1.16-fold, *P*=0.01) and CCL13/MCP4 (Fig. S3D, 1.61-fold, *P*=0.002), as did stimulation with RSV (Fig. S3 E-F) for IFN-α (*P*=0.018) and CCL13/MCP4 (*P*=0.019). These latter two differences were marginal after adjustment (both *P*=0.055, Table 2).

Since adverse clinical outcomes in viral infections are likely in those individuals with the weakest innate anti-viral responses, we examined proportions of males and females whose innate anti-viral responses were below certain lower thresholds. We restricted this analysis to IFN-α and low thresholds were defined as the 15^th^, 20^th^ and 25^th^ percentile of the response determined from the entire population. Results are shown in Fig. 5 and S4. Proportions of males having IFN-α responses to RV-A16, RSV and R848 below each threshold were significantly higher compared to the proportion of females (Fig. 5). Similar trends were observed for response of IFN-α to RV-A1, and CpG-A (Fig. S4), but these did not reach statistical significance.

**Figure 5:**
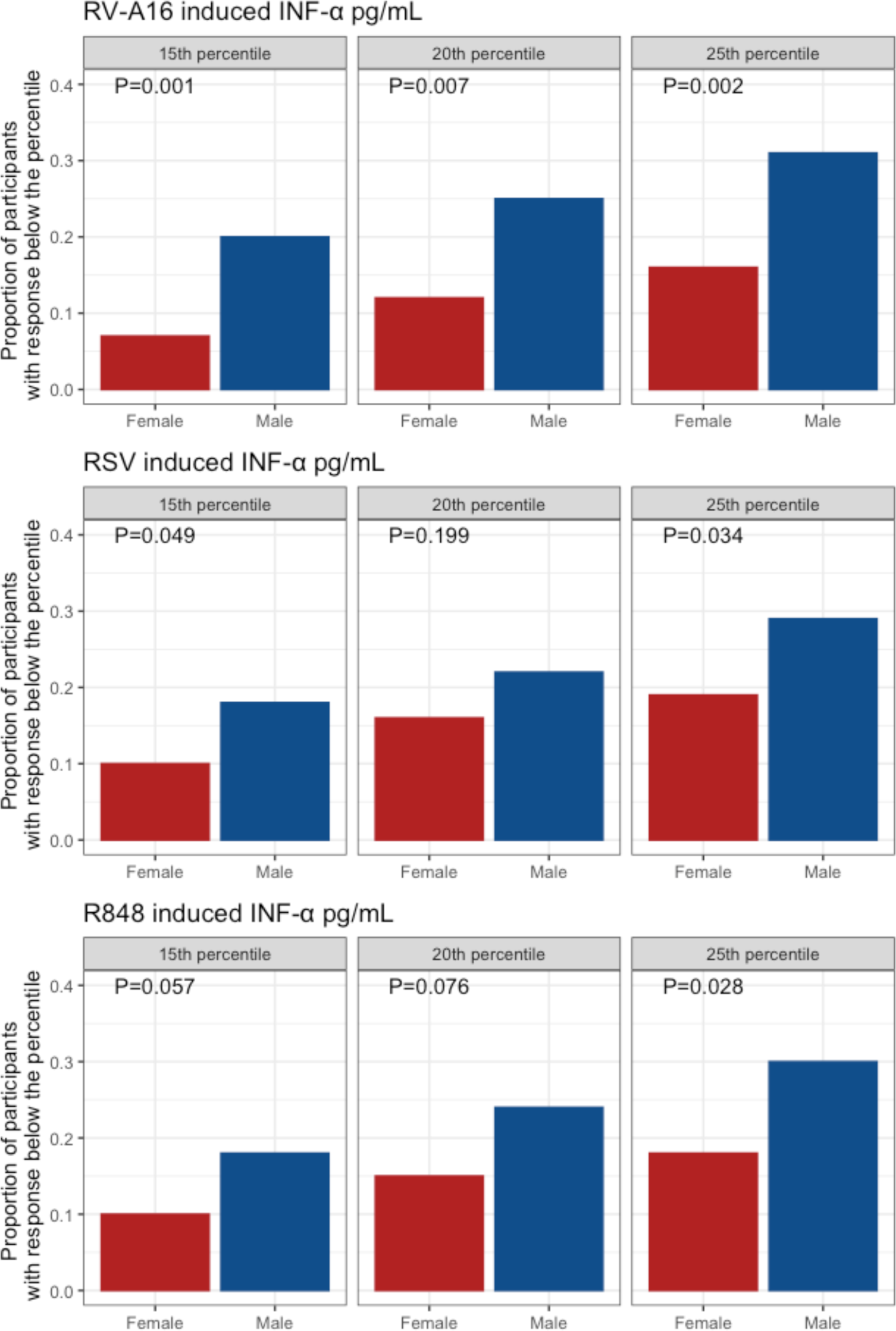
Proportions of males and females with IFN responses to RV-A16, RSV and R848 below the 15^th^, 20^th^ and 25^th^ percentiles of the entire population. *P*-values are derived using chi-squared tests.

### Differences in early-life severe LRTIs between males and females

Having observed diminished innate immune responses to viral stimuli in males, and greater proportions of males with weak innate immune responses, we investigated whether frequencies of early-life LRTIs were also different between sexes in our cohort. We focussed on early life, as this is when innate immune responses will be most important, as the very young will have had little opportunity to develop memory responses. Results are presented in Table S3. Among participants attending follow up at age 16 years who had primary care records available for inspection (n=651), 12.1% of males and 6.6% of females were admitted to hospital with LRTI in the first year of life (*P*=0.017). In the second year of life, 6.1% of males but only 0.75% of females were admitted to hospital with LRTI (*P* <0.001). We observed a similar trend for RSV-positive bronchiolitis, with 4.32% of males hospitalised, compared to 1.64% of females, but the difference between the sexes did not reach formal statistical significance (*P*=0.067).

## Discussion

We hypothesised that the adverse outcomes for males reported in virus infections and especially in SARS-CoV-2 infections, including substantially increased COVID-19-related mortality, ICU admissions, hospital admissions and case identification, may be related to deficient innate immune responses in males relative to females, resulting in increased disease severity in males. Our analysis of induction of three IFNs and four IFN-induced chemokines by five respiratory viruses/viral stimuli in PBMCs sampled from participants in a population-based birth cohort study demonstrated that responses in females were substantially greater than those in males in 32 of 35 cytokine-stimulus pairs. Of these, differences in 14 cytokine-stimulus pairs were statistically significant, of which 9 remained significant after adjustment for multiple testing, all showing increased responses in females. These statistically significant cytokine-stimulus pairs included IFN-α induction in response to each viral stimulus, IFN-β induction in response to RV-A16 and IFN-γ induction in response to RV-A16 and CpG-A.

IFN-α and IFN-β are type I IFNs that are critical mediators of innate anti-viral immune responses, inducing apoptosis of virus-infected cells and inducing over 300 IFN-stimulated genes, many of which have a variety of direct anti-viral activities(20). Through these combined activities, type I IFNs can abort virus replication in virus-infected cells(16).

IFN-γ is the only type II IFN, and it also is very important in promoting innate immune responses principally by activating natural killer (NK) cells which are important in innate immune defense against virus infections, by rapid killing of virus-infected cells(21). IFN-γ also primes other immune cells such as macrophages, to release anti-viral cytokines(22) and to phagocytose infected cells(21). IFN-γ has been shown to suppress mouse coronavirus replication, though this was dependent, in part, on induction of type I IFN secretion(23).

Type I and II IFNs also cooperate and work together to activate macrophages, NK cells, dendritic cells and T cells by enhancing cell activation, antigen presentation, cell trafficking, cell differentiation and proliferation, resulting in markedly enhanced innate and acquired antiviral immune effector function(24). Thus, deficiency in either or both of these IFN types would be expected to markedly increase severity of virus-induced illnesses. We have previously described deficiency in IFN-α(15), IFN-β(15, 16) and IFN-γ(17, 18) in patients with asthma and have reported that patients with asthma have increased susceptibility to respiratory virus infections(19). In addition, recent reports indicate that deficiencies in IFN responses are linked to increased susceptibility to virus-induced exacerbations of chronic obstructive pulmonary disease(25-28). These data therefore support the biological and clinical relevance of our findings.

The deficiencies we observed in IFN responses in males are reflected in a significantly increased incidence of severe LRTIs requiring hospital admissions in the first and second years of life, as well as with a trend for increased incidence of RSV-proven bronchiolitis, a disease with a peak incidence at 4.5 months of age(29). We focused on severe LRTIs requiring hospital admissions in early life, as these are almost exclusively viral in aetiology. The first 2 years of life is a time of life during which innate immune responses will be most important, as infants and toddlers will have had very little experience of viral RTIs and will therefore have little memory/acquired immunity. The fact that we find females produce approximately twice the concentrations of IFN-α and IFN-β in response to RV-A16 and of IFN-α in response to R848, 55% more IFN-α in response to RV-A1 and 71% and 52% more IFN-α and IFN-β in response to CpG-A, than do males, and females have ∼half the number of LRTIs requiring hospital admission in the first year of life, 88% fewer in the second year of life and 62% fewer admissions with RSV-positive bronchiolitis also lends credence to the biological and clinical relevance of our findings in relation to diseases where innate immune responses will be vitally important in protection against adverse outcomes.

SARS-CoV-2 is a virus completely new to mankind, it is therefore even more important to have as strong an innate immune response to this virus as possible, to reduce the likelihood of adverse outcomes. The marked reductions in type I IFN responses to viral stimuli reported herein in males relative to females, with the relationships discussed above with virus-induced exacerbations of lung disease(15-18, 25-28), and with severe viral LRTIs in infants in our cohort, make it almost certain that the deficient IFN responses we report in males will be at least in part responsible for the adverse outcomes to SARS-CoV-2 infection currently being reported in COVID-19 in males(2, 4-10, 12, 13).

A very recent study by Takahashi et al studied sex differences in immune responses during COVID-19 disease in hospitalised patients and claimed that these differences underlie sex differences in COVID-19 disease outcomes(30). This study reported female patients mounted significantly more robust T cell activation than male patients during SARS-CoV-2 infection. They also found that a poor T cell response was associated with worse disease outcome in male patients. This study also reported significantly increased plasma concentrations of IL-8 and IL-18 and numbers of peripheral blood non-classical monocytes in males, with trends to increased SARS-CoV-2 virus load (of magnitude 1-2 Log10 copies/mL, noting that virus load was only measured in the upper respiratory tract and not in the lung) and plasma CXCL-10 concentrations in males. As robust innate interferon responses are well known to drive robust T cell responses during viral infections(24), and deficient innate interferon responses studied *ex vivo* pre-infection are known to relate to increased virus load, increased inflammatory outcomes and worse clinical outcomes upon subsequent *in vivo* respiratory virus infection(31), we suggest all the differential outcomes between sexes identified by Takahashi et al would be a consequence of the deficient innate interferon responses in males, and more robust innate interferon responses in females, that we report herein.

Our study has strengths and limitations. Strengths include the population-based cohort design, so our participants should be representative of the general population, the fact that illness ascertainment was carried out by personal inspection of primary care records to maximise accuracy, that PBMCs were stimulated/infected with the most common respiratory viruses and with ligands of TLRs that are representative of RNA respiratory viruses such as SARS-CoV-2, and that infections/stimulations were carried out using a single batch of each virus/stimulus and conducted by a single highly experienced individual, so any variability in response will be participant-related and not a result of technical variability. Limitations include that PBMCs were not directly infected/stimulated with SARS-CoV-2. This was not possible as our participants’ PBMCs were infected/stimulated before SARS-CoV-2 existed as a human pathogen. However, we did study ligands (R848 and CpG-A) to the endosomal expressed TLRs 7/8/9, which are clearly engaged by positive strand RNA viruses such SARS-CoV-2, which require endosomal processing as part of viral entry into cells(32). Furthermore, consistency of findings across all viral stimuli and responses suggests that our findings should apply to all respiratory viruses including SARS-CoV-2.

We were not able to assess the importance of induction of the type III IFNs, IFN-λs 1-3, which are very important in innate antiviral responses against respiratory viruses(31, 33-35), as the type III IFNs were not significantly induced by any infection/stimulus in the PBMCs that we studied (data available on request). This is likely because respiratory epithelial cells are the main source of type III IFNs, and the cells within PBMCs that are capable of producing type III IFNs when studied fresh(35) likely succumbed as a result of freeze/thawing.

Our findings have important therapeutic implications. IFN-α and IFN-β have both been shown to inhibit replication of SARS-CoV-2(36, 37). A very recent report indicates that early subcutaneous IFN-β administered up to three times weekly (with ribavirin) in the first seven days after diagnosis of COVID-19 was associated with faster virus clearance and clinical improvement and with shorter hospital stays(38). It is likely that most of this clinical benefit resulted from IFN-β rather than ribavirin(39), but ongoing clinical trials (more than 20 trials investigating IFN-α or IFN-β in COVID-19 are currently registered on clinical trial registries) will determine whether and to what degree type I IFN therapies are effective.

Since IFN production by virus-infected respiratory cells is likely critical to innate anti-viral immunity against SARS-CoV, therapeutic use of agents that boost innate IFN induction by virus infections may be efficacious in treatment of COVID-19. We reported 10 years ago that azithromycin doubles IFN-β and IFN-λ production from virus-infected human bronchial epithelial cells *in vitro*(40). Erythromycin and telithromycin did not(40). Innate IFN-induction by azithromycin in virus-infected human lung cells has been confirmed in three subsequent studies(41-43), one of which confirmed this property was variable among 225 novel macrolides studied, as some macrolides induced innate IFNs (up to ∼5-fold), while others did not(43). Thus this innate IFN-inducing property is not a property of all macrolides, but is reproducibly a property of azithromycin(40-43). This is supported by a recent report that among 1,520 clinically approved compounds from a chemical library screened for anti-viral activity against SARS-CoV-2, only two had clear dose-responsive replication-inhibition curves with identified EC50 concentrations in the 1-10μM range. These two compounds were azithromycin and hydroxychloroquine, and they also had EC90 concentrations of ∼10μM and ∼15μM respectively, with azithromycin achieving ∼100% inhibition of SARS-CoV-2 replication at ∼15μM(44). Azithromycin treatment of COVID-19 patients (especially early in the disease course when patients progress from mild to moderate disease severity) in order to boost IFN production by respiratory epithelial cells when infected with SARS-CoV-2, may therefore be effective, especially in males who demonstrably have deficient innate IFN responses when compared to females. This deduction is indirectly supported by high-quality randomised, double-blind, placebo-controlled clinical trial evidence that azithromycin is effective in preventing progression to severe lower respiratory tract illnesses (which are initiated by viral infections) in preschool children(45) and in prevention of asthma exacerbations (the great majority of which are caused by respiratory viruses) in uncontrolled moderate/severe asthma(46). Ongoing studies (more than 50 clinical trials investigating azithromycin in COVID-19 are currently on trial registries) will determine the effectiveness of azithromycin, and we would suggest that post-hoc analyses include stratification by sex.

Our findings have important implications for prevention strategies, which are best implemented when high-risk populations can be identified. Respiratory viruses, including SARS-CoV-2, are initially encountered by the nasal epithelium where infection is initiated. An adequate anti-viral response can contain the virus in the upper airway, with cold-like symptoms, and a diminished response can see the infection spreading to the lower airways to induce severe LRTI. Our data support a preventive strategy, particularly among males, which involves targeting the innate immune system with ‘immune training’ agents to boost resistance to primary infection with SARS-CoV-2 and enhance the capacity to control the intensity of airways’ inflammatory responses (such as, for example, OM-85(47), which has been shown to effectively and safely reduce winter hospitalizations in COPD(48), severe bronchitis among residents in care-homes for elderly(49), and LRTIs in children(50)). Effective prophylactic therapy will be crucially important while we wait for a vaccine.

Our findings also have important implications for a hotly debated topic, namely whether “man flu” actually exists or not(51). Here we provide a biological basis to explain why males would be expected to suffer more than females when infected by respiratory viruses.

In conclusion, high morbidity and mortality from respiratory viruses including COVID-19 in males is likely explained by impaired innate anti-viral immune in males compared to females. Males may preferentially benefit from therapies that boost innate anti-viral immune responses to viruses, both for treatment and prevention.

## Materials and Methods

Detailed methods are presented in the Supplementary Appendix.

### Study design, setting, participants and data sources

The study subjects were participants in the Manchester Asthma and Allergy Study, a population-based birth cohort study(52). The study was approved by the research ethics committee; we obtained written informed consent. PBMCs were collected from subjects at age 16 years and cryopreserved. We used male or female sex as assigned at birth.

### Cell stimulations and cytokine measurement

The PBMCs were thawed on the day of stimulation, counted and had viability checked as described(53, 54). Cells were distributed in 96-well round bottom plates at 2^*^10^5^ cells/well and were stimulated with two rhinoviruses (RVs), RV-A16 and RV-A1, and respiratory syncytial virus (RSV). We also used two viral mimics (which mimic infection with SARS-CoV-2), the Toll-like receptor (TLR)-7/8 ligand resiquimod (R848) and the TLR-9 ligand class A CpG oligonucleotide (CpG-A), both at 1μM concentrations (Invivogen). Concentrations of IFN-α, IFN-β, IFN-γ and four IFN-induced chemokines CXCL10/IP-10, CCL2/MCP1, CCL4/MIP-1β and CCL13/MCP4 were measured in supernatants 24h post-stimulation, using the Meso Scale Discovery® kits as previously described(53, 54).

### Clinical outcomes: Lower respiratory tract infections (LRTI)

We extracted data on severe LRTIs requiring hospital admissions in the first and second year of life from electronic and paper-based primary care medical records; age in days at the time of each event was documented to provide an accurate account of each episode(54).

### Statistical analysis

#### Quality control

Prior to analyses, data were pre-processed according to the pipeline described in our previous study(53), to exclude samples with low viability and/or no response of any cytokine to any stimulus.

#### Analysis

We expressed the induction of IFNs and IFN-induced chemokines as raw values (in pg/mL) and fold-induction over medium controls. The data were summarised as median and interquartile range (IQR).

Univariate comparisons between groups were performed using the Wilcoxon rank sum test (2-tailed). Benjamini-Hochberg correction was applied to account for multiple testing(55). Proportions of males and females with IFN responses to viral stimuli below the 15^th^, 20^th^ and 25^th^ percentiles of the entire population were compared using chi-squared tests. Associations with *P*<0.05 were considered significant.

## Data availability

The data that support the findings of this study are available from the corresponding author upon reasonable request.

## Author contributions

All authors contributed to the writing of the manuscript and have approved the final version for publication. E.R., S.F., L.L., R.H., J.C., M.R. and M.E. performed the data analyses and E.R., S.F., graph and table production. E.R. performed all cell infections/stimulations and all cytokine measurements. S.H. performed primary care record inspection. A.S. coordinated the clinical study, A.C. and S.J. lead the study design, supervision and interpretation of the studies.

A.S., A.C. and S.J. are responsible for the overall content as guarantors. The guarantors accept full responsibility for the work, the conduct of the study, had access to the data, and controlled the decision to publish. The corresponding author attests that all listed authors meet authorship criteria and that no others meeting the criteria have been omitted.

